# Occurrence and Timing of Subsequent SARS-CoV-2 RT-PCR Positivity Among Initially Negative Patients

**DOI:** 10.1101/2020.05.03.20089151

**Authors:** Dustin R. Long, Saurabh Gombar, Catherine A. Hogan, Alexander L. Greninger, Vikas O’Reilly Shah, Chloe Bryson-Cahn, Bryan Stevens, Arjun Rustagi, Keith R. Jerome, Christina S. Kong, James Zehnder, Nigam H. Shah, Noel S. Weiss, Benjamin A. Pinsky, Jacob Sunshine

**Affiliations:** University of Washington School of Medicine, Department of Anesthesiology & Pain Medicine, Division of Critical Care Medicine, Seattle, WA, USA; Department of Pathology, Stanford University School of Medicine, Stanford, CA, USA; Clinical Virology Laboratory, Stanford Health Care, Stanford, CA, USA; University of Washington School of Medicine, Department of Laboratory Medicine, Seattle, WA, USA; Fred Hutchinson Cancer Research Center, Vaccine and Infectious Disease Division, Seattle, WA, USA; University of Washington School of Medicine, Department of Anesthesiology & Pain Medicine, Seattle, WA, USA; University of Washington School of Medicine, Division of Allergy and Infectious Diseases, Seattle, WA, USA; Division of Infectious Diseases and Geographic Medicine, Department of Medicine, Stanford University School of Medicine, Stanford, CA, USA; Center for Biomedical Informatics Research, Stanford University, Stanford, CA, USA; University of Washington School of Public Health, Department of Epidemiology, Seattle, WA, USA

## Abstract

**Background:** SARS-CoV-2 reverse transcriptase polymerase chain reaction (RT-PCR) testing remains the cornerstone of laboratory-based identification of patients with COVID-19. As the availability and speed of SARS-CoV-2 testing platforms improve, results are increasingly relied upon to inform critical decisions related to therapy, use of personal protective equipment, and workforce readiness. However, early reports of RT-PCR test performance have left clinicians and the public with concerns regarding the reliability of this predominant testing modality and the interpretation of negative results. In this work, two independent research teams report the frequency of discordant SARS-CoV-2 test results among initially negative, repeatedly tested patients in regions of the United States with early community transmission and access to testing.

**Methods:** All patients at the University of Washington (UW) and Stanford Health Care undergoing initial testing by nasopharyngeal (NP) swab between March 2nd and April 7th, 2020 were included. SARS-CoV-2 RT-PCR was performed targeting the *N, RdRp, S*, and *E* genes and ORF1ab, using a combination of Emergency Use Authorization laboratory-developed tests and commercial assays. Results through April 14th were extracted to allow for a complete 7-day observation period and an additional day for reporting.

**Results:** A total of 23,126 SARS-CoV-2 RT-PCR tests (10,583 UW, 12,543 Stanford) were performed in 20,912 eligible patients (8,977 UW, 11,935 Stanford) undergoing initial testing by NP swab; 626 initially test-negative patients were re-tested within 7 days. Among this group, repeat testing within 7 days yielded a positive result in 3.5% (4.3% UW, 2.8% Stanford) of cases, suggesting an initial false negative RT-PCR result; the majority (96.5%) of patients with an initial negative result who warranted reevaluation for any reason remained negative on all subsequent tests performed within this window.

**Conclusions:** Two independent research teams report the similar finding that, among initially negative patients subjected to repeat SARS-CoV-2 RT-PCR testing, the occurrence of a newly positive result within 7 days is uncommon. These observations suggest that false negative results at the time of initial presentation do occur, but potentially at a lower frequency than is currently believed. Although it is not possible to infer the clinical sensitivity of NP SARS-CoV-2 RT-PCR testing using these data, they may be used in combination with other reports to guide the use and interpretation of this common testing modality.

## Introduction

Severe acute respiratory syndrome coronavirus-2 (SARS-CoV-2) is the etiologic agent of coronavirus disease 2019 (COVID-19). Accurate detection of the virus is essential to strategies endorsed by the Centers for Disease Control and World Health Organization. As the availability and speed of SARS-CoV-2 testing platforms improve, results of these tests are increasingly relied upon to inform critical decisions related to therapeutic intervention, use of personal protective equipment, patient isolation, and workforce readiness. While the analytic performance of SARS-CoV-2 reverse transcriptase polymerase chain reaction (RT-PCR) tests are well described[1], clinical performance is impacted by several factors that are difficult to measure, such as low levels of shedding during incubation and early infection[2], variability in the site of specimen acquisition[3,4], and sufficiency of sample collected. In addition, early reports and characterizations in the press have left the medical community and general public with concerns about the reliability of SARS-CoV-2 RT-PCR testing and the interpretation of negative results. Data characterizing the scope of false negative results observed in the context of current US testing practices are needed to guide clinical protocols and inform the public, but are lacking.

The initial US introduction of COVID-19 through Washington State [5] followed closely by Northern California[6], combined with the early availability of SARS-CoV-2 testing in both regions[7,8], provides an opportunity to evaluate clinical test performance in a population of repeatedly tested patients. In this study, utilizing data from two independent healthcare systems and analyzed by separate research teams, we report the frequency of discordant SARS-CoV-2 RT-PCR results among initially test-negative individuals who were subsequently retested within 7 days.

## Methods

### Common Study Methods

All patients at both sites undergoing initial testing for COVID-19 by SARS-CoV-2 RT-PCR of a nasopharyngeal (NP) swab between March 2nd and April 7th, 2020 were included. Test results through April 14th were extracted to allow for a complete 7-day observation period and an additional day for result reporting. Data on cycle threshold (Ct) values were extracted for each test, and are interpreted as inversely proportional to the viral load level present in the sample. Inconclusive RT-PCR test results (i.e., only 1 of 2 SARS-CoV-2 target genes amplified) were treated as positive in accordance with institutional clinical guidelines.

### UW Methods

The UW Virology clinical laboratory serves as the primary testing center for a broad region in the US Pacific Northwest, processing over 60% of all SARS-CoV-2 tests for Washington State during the time period examined. In order to ensure consistency of clinical data and compliance with patient privacy policies, analysis was limited to adult patients having an established affiliation with UW Medicine. Encounters spanning multiple facilities (e.g. outpatient, hospital, and drive-through testing locations) were linked using an unambiguous identifier common to all sites. UW guidelines over the study period for testing included the following: all patients who exhibited one or more symptoms of COVID-19 at the time of initial testing per institutional protocol, which involved new symptoms of acute respiratory infection (e.g., fever, cough, shortness of breath, myalgias, rhinorrhea, sore throat, anosmia, ageusia), combined with pertinent risk factors (occupation, age, chronic disease status, immunosuppression, contact with confirmed COVID-19 cases, pregnancy, housing stability, exposure to high-risk facilities or inpatient admission) or based on clinical judgement; starting March 30th, 2020, universal pre-operative screening for all asymptomatic surgical cases. Nasopharyngeal samples were collected according to standard UW protocol (**Supplement 1**). UW testing platforms included a laboratory-developed test (LDT) two-target/two-control assay modified from the CDC (target genes *N1, N2)* operating under a Washington State emergency use authorization[7]; Panther Fusion SARS-CoV-2 assay (Hologic, Marlborough, MA, target genes two conserved regions of ORF1ab); Roche RT-PCR (Basel, Switzerland, target *E* gene); DiaSorin (Saluggia, Italy, target ORF1ab and *S* genes). The University of Washington Institutional Review Board determined this study to be exempt (STUDY00009931).

### Stanford Methods

The Stanford Health Care (SHC) Clinical Virology Laboratory is based in northern California and performed SARS-CoV-2 testing on both adult and pediatric populations. Approximately 2/3 of the samples were from Stanford Medicine facilities and 1/3 were from medical facilities in northern California with the greatest concentration coming from facilities in San Mateo and Santa Clara counties. Testing was performed using one of two assays: 1) SHC Emergency Use Authorization LDT^6^ or 2) Panther Fusion SARS-CoV-2 assay. This study received approval by the Stanford Institutional Review Board (Protocol #48973) and individual consent was not required.

## Results

A total of 23,126 SARS-CoV-2 RT-PCR tests (10,583 UW, 12,543 Stanford) were performed in 20,912 eligible patients (8,977 UW, 11,935 Stanford) undergoing initial testing by NP swab between March 2nd and April 7th, 2020. Initial results for 91% (90.7% UW, 91.2% Stanford) of patients were negative (**Figure 1A**). Characteristics of initially negative patients are shown in **Supplemental Table 1**. The majority of these patients (95.9% UW, 97.4% Stanford) did not undergo repeat testing within 7 days and did not require subsequent evaluation in the form of outpatient, emergency department, or inpatient encounters (**Supplemental Table 1**). Results of other viral respiratory tests were available for UW patients and several negatively retested patients were ultimately diagnosed with other viral respiratory illnesses, most commonly influenza A, rhinovirus, and RSV (**Supplemental Table 1**). However, a small proportion (4.1% UW, 2.6% Stanford) underwent repeat testing within this window despite an initial negative result (**Figure 1A**). Among those requiring reevaluation, 96.5% (95.9% UW, 97.2% Stanford) remained negative on all repeat tests performed within 7 days.

**Figure 1.**
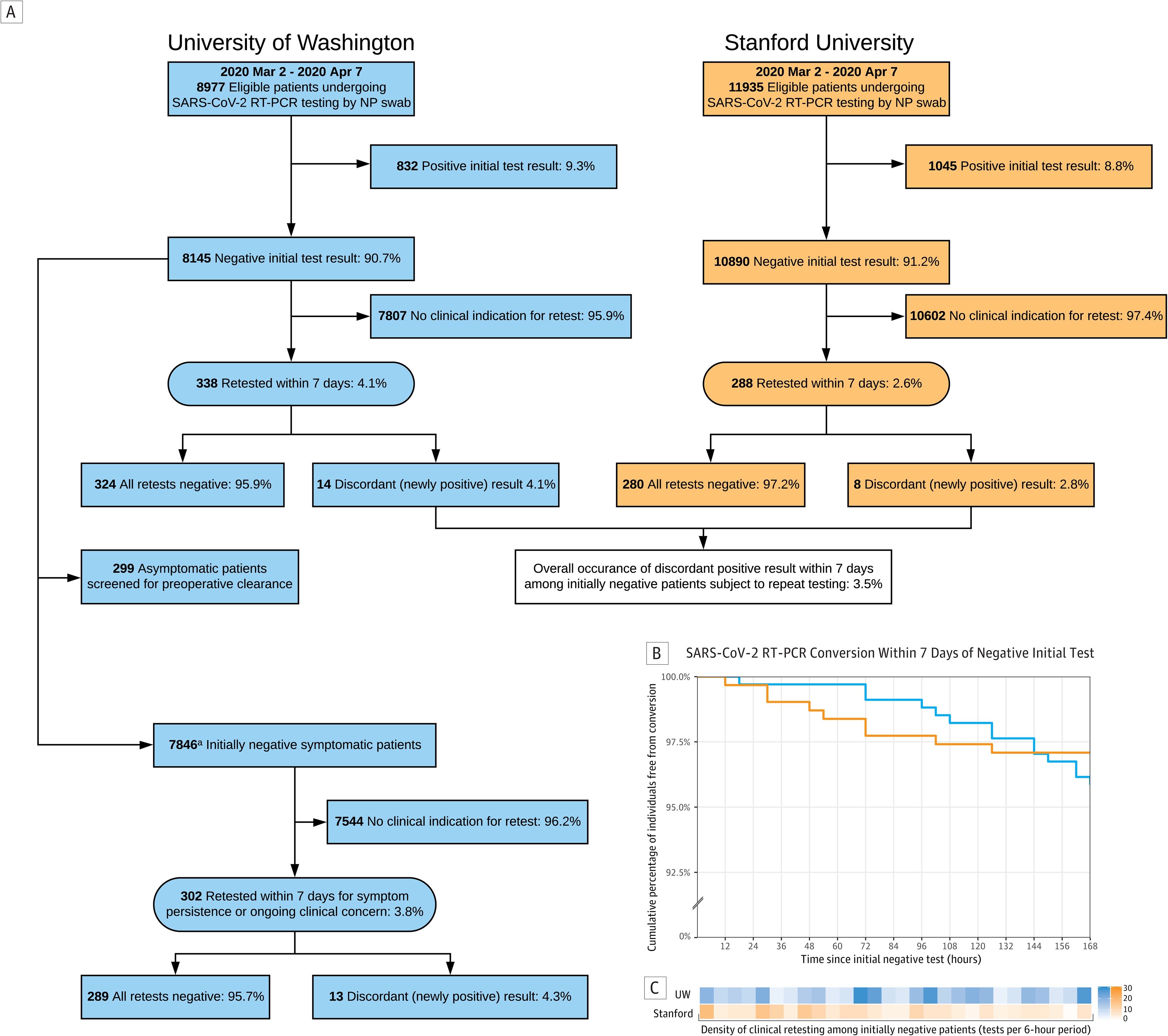
Identification of patients initially testing negative for SARS-CoV-2 and outcomes of repeat testing. A. The primary measure was the occurrence of a discordant (newly positive) result within 7 days. ^a^Subgroup analysis excluding asymptomatic patients screened for surgical clearance at the University of Washington yielded similar results. B. Among patients initially testing negative for SARS-CoV-2 by RT-PCR of a nasopharyngeal swab, over 95% of patients at both UW (blue) and Stanford (orange) subjected to retesting remained negative on subsequent tests performed within 7 days. C. Retesting of initially negative individuals occurred at varied intervals across the 7-day period of observation.

It was observed that 3.5% (4.1% UW, 2.8% Stanford) of patients subjected to retesting on clinical grounds within 7 days were subsequently found to be positive during this period, suggesting a false negative initial result. The timing of clinical retesting and conversion among these patients is shown by site in **Figure 1B and Figure 1C**, respectively. The clinical contexts and testing parameters of the 22 patients with discordant results are summarized in **Supplemental Table 2**. In this group, the mean interval between initial negative test and first positive retest was 4.0 days (SD 2.0). RT-PCR cycle threshold values of newly positive results averaged 28.5 (SD 8.0), consistent with lower viral RNA loads.

At UW, the use of standardized testing algorithms enabled subgroup analysis based on testing indication (**Figure 1A**). A total of 299 asymptomatic individuals who were tested as part of universal screening for preoperative clearance were excluded, leaving 7,846 symptomatic individuals who tested negative at the time of initial presentation for analysis. Of the 302 individuals in this group with persistent or worsening symptoms warranting additional testing within 7 days, 4.3% converted from negative to positive and 95.7% remained negative on all subsequent SARS-CoV-2 tests performed within this window.

## Discussion

In this report, two independent research teams describe that, among patients initially testing negative by SARS-CoV-2 RT-PCR of a NP swab, repeat testing within 7 days yielded a positive result in 3.5% of cases; the majority (96.5%) of those warranting additional testing for any reason remained negative on all subsequent tests within this window. Among the subgroup of UW patients confirmed to have symptoms prior to an initial negative result who were retested for persistent or worsening symptoms, a similar proportion (4.3%) were subsequently found to be positive within 7 days. These observations suggest that false negative NP SARS-CoV-2 RT-PCR results do occur, but potentially at a lower frequency than is currently believed.

Results from each research group have limitations. Neither team is able to calculate a true clinical sensitivity or false negative proportion due to the absence of retesting in all initially negative patients and the lack of a gold standard confirmatory mechanism. Additionally, it cannot be ruled out that some discordant test results in this cohort may be due to newly acquired infection. By limiting the scope of retesting considered to a 7-day period, the likelihood of this scenario is minimized, but not eliminated. Finally, we were unable to ascertain the disease status of the individuals who initially tested negative for COVID-19 but did not undergo repeat testing; in most cases this likely reflects the absence of an indication for retesting (e.g. alternative diagnosis or resolution of symptoms), but could also be the result of limited access to care.

The intention of this report is not to definitively quantify the clinical performance of NP SARS-CoV-2 RT-PCR testing, which will likely require orthogonal approaches such as serology. Rather, by characterizing the experience of two large US health systems on the short-term occurrence of newly positive SARS-CoV-2 results among initially test-negative patients, we provide data on a topic of practical significance that should be used in combination with other reports to guide the use and interpretation of this common testing modality.

## Data Availability

NA

## Acknowledgments

We would like to thank Christine Fong and the Center for Perioperative & Pain initiatives in Quality Safety Outcome (PPiQSO) at the University of Washington, Seattle for assistance with the data extract analyzed in the present work.

